# Classification of Hyper-scale Multimodal Imaging Datasets

**DOI:** 10.1101/2023.01.03.23284138

**Authors:** Craig MacFadyen, Ajay Duraiswamy, David Harris-Birtill

## Abstract

Algorithms that can classify hyper-scale multi-modal datasets, comprising of millions of images, into constituent modality types can help researchers quickly retrieve and classify diagnostic imaging data, accelerating clinical outcomes. This research aims to demonstrate that a deep neural network that is trained on a hyper-scale dataset (4.5 million images) composed of heterogeneous multi-modal data, can be used to obtain significant modality classification accuracy (96%). By combining 102 medical imaging datasets, a dataset of 4.5 million images was created. A ResNet-50, ResNet-18 and VGG16 were trained to classify these images by the imaging modality used to capture them (Computed Tomography (CT), Magnetic Resonance Imaging (MRI), Positron Emission Tomography (PET), and X-ray) across many body locations. The classification accuracy of the models was then tested on unseen data.

The best performing model achieved classification accuracy of 96% on unseen data. The model achieved a balanced accuracy of 86%.

This research shows it is possible to train Deep Learning (DL) Convolutional Neural Networks (CNNs) with hyper-scale multimodal data-sets, composed of millions of images. The trained model can be used to classify images by modality, with the best performing model achieving a classification accuracy of 96%. Such models can find use in real-world applications with volumes of image data in the hyper-scale range, such as medical imaging repositories, or national healthcare institutions. Further research can expand this classification capability to include 3D-scans.

## 1 Introduction

With the proliferation of deep neural networks trained on heterogenous multimodal data to detect and predict diseases, there has been an explosion in the volume of diagnostic medical imaging data [1]. Clinicians often order multiple scans of the same patient in different modalities to gather evidence to make improved diagnosis/prognosis [2]. Algorithms that can accurately classify a large hetergogenous dataset into its constituent modalities can be beneficial to researchers and clinicians, allowing them to automatically segment a particular type of modality for retrieval, archival, data balancing, and diagnostic purposes. Manual methods for classifying medical images are typically error-prone unless done by costly domain experts [3].

This paper outlines an deep neural network that accurately classifies a hyper-scale (4.5 million images), mixed-modality dataset into constituent modalities. The developed approach has significant benefit potential for researchers, clinicians, and imaging archives by helping effectively and efficiently classify diagnostic imaging data, in the magnitude of real-world volumes. While classification of hyperscale datasets have been attempted in other areas, such as Earth-science[4], including studies of plankton and marine snow [5], and XYZ, the proposed approach is novel in the field of classification of medical imaging modalities. This study aims to stimulate other hyper-scale projects in this area.

Multiple open-access data sets were used to build the hyper-scale multimodal dataset of 4.5 million images from sources such as The Cancer Imaging Archive [16], Stanford ML Group[17] the largest of which contains 262,000 chest X-ray images, and Kaggle [18] host labelled datasets.

The models trained on this hyper-scale multimodal dataset were a ResNet-18,23 ResNet-50 and a VGG16. When these models were tested for classification accuracy, these results are in the high 90%’s across the train, validate and test sets which shows that the models are able to classify with significant accuracy. Table 1 shows the accuracy and balanced accuracy of each of the models on the test set. Figure 6 shows the confusion matrix for the ResNet18, the best performing of the models tested in this study. The confusion matrix shows that the model demonstrates significant classification performance (96%+) on classifying CT, MR and PET modalities.

**Table 1:** Dataset sizes vs Performance

| Study | Dataset | Classifier |
| --- | --- | --- |
|  | Magnitude | Accuracy |
| Chiang et al.[25] | $10^3$ | $> 99.5\%$ |
| Cheng et al.[26] | $10^2$ | $> 89.6\%$ |
| Yu et al.[27] | $10^3$ | 70% |
| Sevakula et al.[28] | $10^3$ | 99.45% |
| Trenta et al.[24] | $10^3$ | 100% |

### 1.1 Previous Literature

A number of research articles focus on deep learning systems to classify modalities in diagnostic imaging data. However, to the best of our knowledge,there have not been any examples of a system that combines medical imaging datasets at the hyper-scale (millions of images) level to perform modality classification.

Approaches to classifying medical imaging data by modality primarily take two forms (1) hand-crafted features, and (2) Deep Learning.

The early approaches were based on hand-crafted features, such as picking a specific texture and colour[19], SIFT descriptors[20], bag-of-colours[21] and then using SVM [22], KNN [23] as the classifier[24]. These approaches were limited by the choice of features, and limited accuracy[3]. Further, typically high computational costs inherently limit the size of the datasets used.

A number of approaches using deep learning classifiers are seen in literature. However, all approaches reviewed were seen to be utilising a limited dataset volumes with sizes in the lower order of magnitude, typically hundreds to thousands (10^2^ *−*10^3^) of images. Therefore, real-world classification performance of these algorithms when operated on typical image-repository scales of millions of images is unknown.

Chiang et al. use a dataset of 2,878 images to train a CNN classifier on 4 modalities[25], Abdominal CT, Brain CT, Lumbar Spine MRI, and Brain MRI, achieving an average validation accuracy of > 99.5%. Cheng et al. use a cascaded CNN to classify a bimodal dataset, comprised of MRI and PET images[26]. Using a dataset in the order of 10^2^ images, they achieved a classification accuracy of 89.6%. Yu et al. use a DNN, and a dataset from the ImageCLEF database, comprising of 2,901 training and 2,582 test images to demonstrate a best classification accuracy of 70%[27]. Sevakula et al. use transfer learning to compare performance of seven DCNNs[28]. Using a curated dataset of 5,500 images from the Open-i Biomedical Image Search Engine, they achieve a best classification accuracy of 99.45% on the Inception-V3 network. Finally, Trenta et al. use a dataset comprised of 8,500 slices and a test set of 1,320 slices (split across 5 classes), and transfer learning techniques to achieve an overall accuracy of upto 100% on specific modalities, on their pre-trained VGGNet implementation[24].

Summarising classification performance figures reported in extant literature:

To summarise, two thing are evident, (1) deep learning learning models present several advantages over handcrafted, feature driven models, and (2) it is seen that the largest of the datasets in the literature reviewed is in the order of 10^3^ images. Given that image repositories are now typically in the hyper-scale order, and growing rapidly, a suitably trained CNN capable of handling hyper-scale datasets is required.

## 2 Materials & Methods

### 2.1 Data

In total, 102 datasets were downloaded and combined to form an hyper-scale image dataset of 4.5 million images. The full list of datasets with citations is provided in Appendix A. Four modalities were selected as targets for the classification task; CT, MRI, X-ray and PET. Other modalities (e.g. ultrasound) were excluded from this study because of a lack of appreciable volumes of data. The main source of this data was the Cancer Imaging Archive (TCIA) [16]. The Cancer Imaging Archive provides a REST API that allows for programmatic retrieval of images which allowed data to be downloaded and combined easily, and in a reproducible way. However, because the Cancer Imaging Archive’s main purpose is to host datasets relating to cancer research it was important to seek out some extra datasets to augment the data TCIA provides. The full list of datasets can be found in Appendix A.

This project was approved by the University of St Andrews University Teaching and Research Ethics Committee (UTREC), approval code CS15171.

### 2.2 Train-Validate-Test Split

The downloaded data was split into three separate parts - train, validate and test. The train set was used to train the model, the validate set was used to evaluate the models between training runs, and the test set was used once to evaluate the final trained models. It was important to create the splits at the dataset level to prevent data-leakage. That is, all the images from a dataset were placed in the same split. Scans of the same patient in the same modality are likely to be similar, so if there is an image of the same patient in the train and test set then the test set does not contain completely unseen data. Putting each dataset into one of train, validate or test prevents this data leakage. Splitting the datasets like this also helps achieve the goal of demonstrating generalisation across datasets, because no dataset in the train set is represented in the test set.

The train-validate-test split was created manually to ensure as even a spread as possible of images for each modality and location in each split. The manual split ensured that there are at least two locations for each modality in each of the train, validate and test split. The main difficulty for this was X-rays, because in the TCIA datasets most X-rays are mammograms. This meant the non-TCIA datasets had to be carefully split. Again, Appendix A shows the split each dataset was placed in. Figure 3c shows the number of images in the train, validate and test set. TCIA hosts many CT and MR datasets and some of these datasets are very large. For example, the CT Colonography dataset [29] has more than 900,000 CT images, which is more than the total number of X-ray images across all datasets used in this study. To ensure the other modalities were not completely dwarfed by these datasets, a maximum of 50,000 CT images and 100,000 MR images was taken from each individual dataset. The images were selected in the order given by TCIA. This selection method was not applied to the images from sources other than TCIA. After imbalance correction, the total number of images in the dataset were 6,433,838 (6.4 million images), with a spilt of 4,104,184 in training, 936,347 in test, and 1,393,307 in validate datasets.

**Figure 1:**
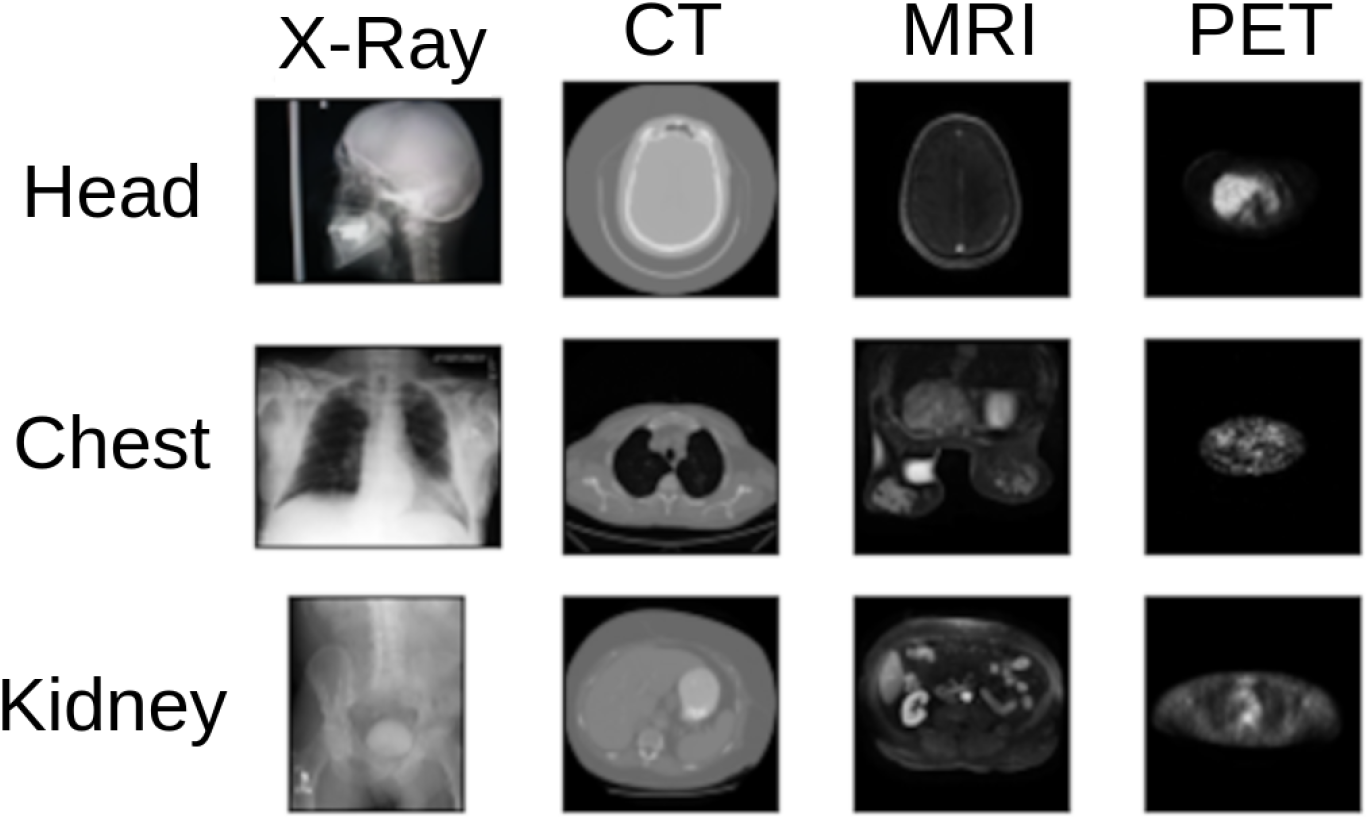
Visualisation of a spread of images from different locations in different modalities. Different modalities use different kinds of radiation, and these are absorbed to varying degrees by tissue in the human body. This leads to the same tissue looking different in each modality. Examples of modalities showing variation of the same tissue: [6] [7] [8] [9] [10] [11] [12] [13] [14] [14] [15]

**Figure 2:**
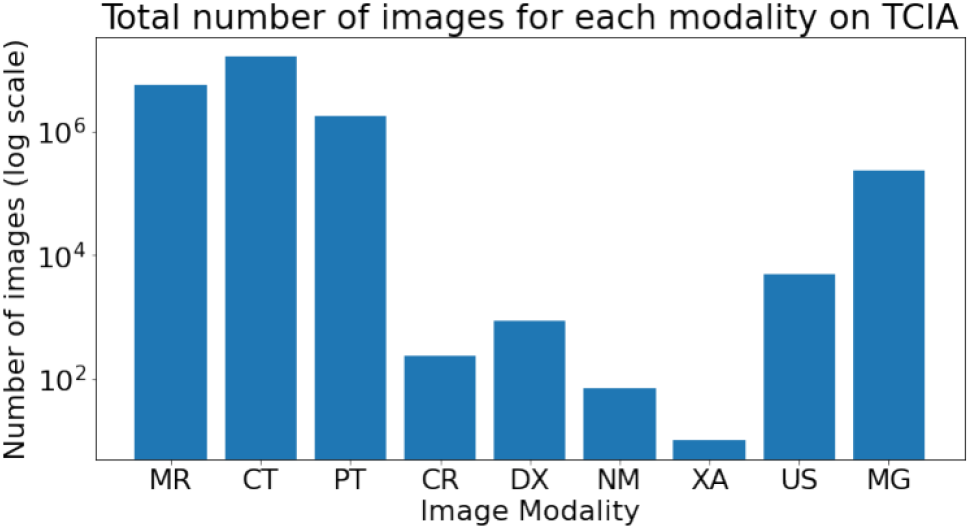
XXX

**Figure 3:**
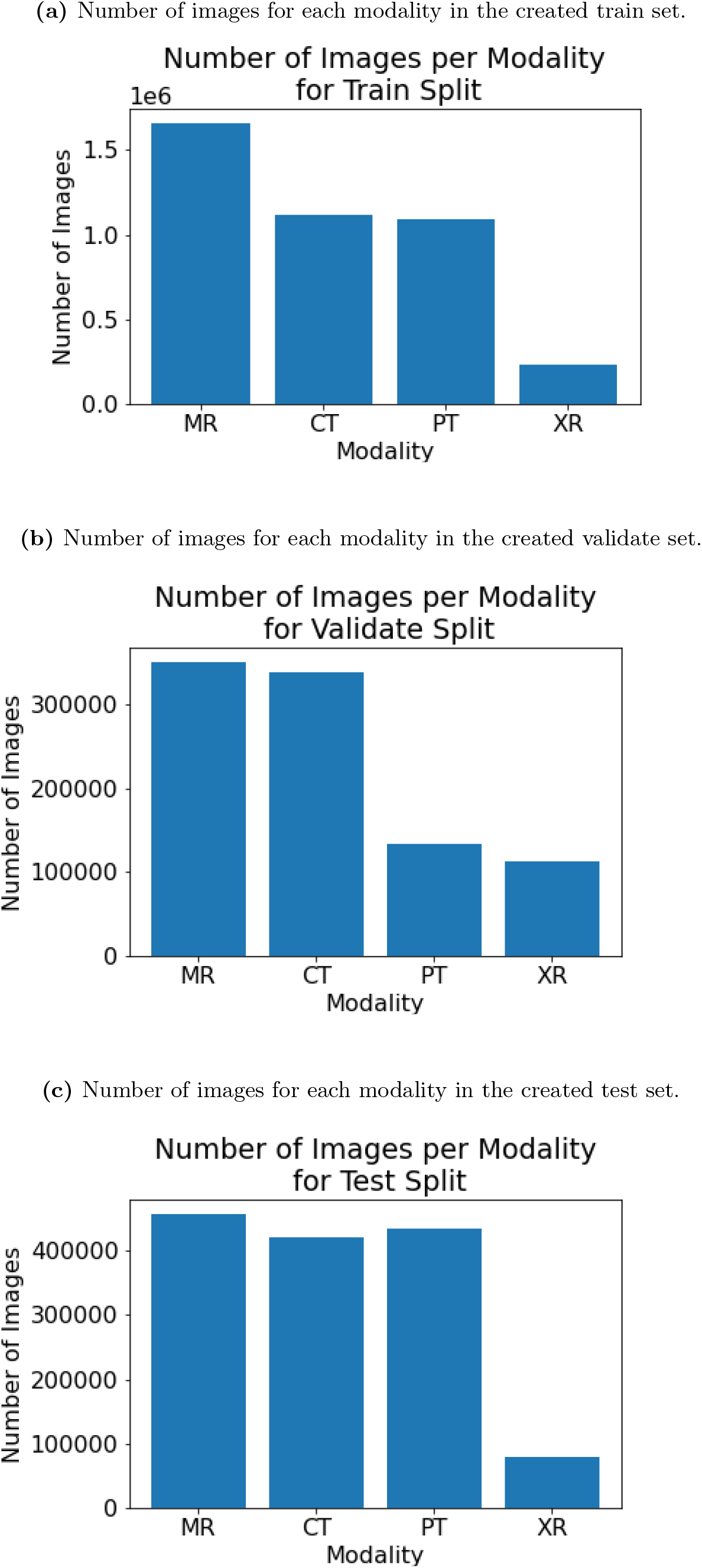
Figures showing the number of images for each modality in the created splits: a) train, b) validate and c) test. Note that each graph has a different scale, the purpose is to show the ratios of each class are similar. There are 73 datasets in the train set, 13 in the validate set and 16 in the test set.

### 2.3 Preprocessing

In order for 2D and 3D scans to be used in the same study, the 3D scans (CT, MR and PT) were treated as a collection of 2D images. These images are sometimes referred to as slices. The images were resized to 224 × 224 and rescaled between 0 and 1. Each image was rescaled using min-max normalisation with the maximum and minimum values being the highest and lowest pixel values present within the image.

### 2.4 Network Architecture and Training

The models trained on this dataset were a ResNet-18 [30], ResNet-50 and a VGG16 [31]. The code used was adapted from PyTorch’s hosted versions of these models[32]. Changes were made to the channel depth of the input layer, from three channels to one channel (grayscale). These three models were chosen because they have all been shown to perform well when trained with large quantities of data on the ImageNet dataset [30, 31]. The code created as part of this research is open-source and hosted online at Github [33].

All models were trained for 10 epochs with a batch size of 128. The training set contained 2,954,097 (2.9*x*10^6^) samples and the validate set contained 704,685 samples. The models were optimised using stochastic gradient descent, with a learning rate of 0.1 that was divided by 10 every time the loss plateaued, a momentum of 0.9 and an L2 weight decay penalty of 0.005. The models were trained on a machine with an Intel(R) Xeon(R) CPU E5-1650 v4 @3.60GHz with 6 physical cores (12 threads), 250GB of RAM and two Nvidia GeForce GTX 1080Tis.

## 2 Results & Discussion

### 3.1 Training and Validation Accuracy

Figures 4, shows the training and validation accuracy curves for the ResNet50, ResNet18 and VGG16 models. The small gap between the training and validation accuracies suggests that the models are not overfitting. Figure 5 shows the time it took to train the models over the 10 epochs.

**Figure 4:**
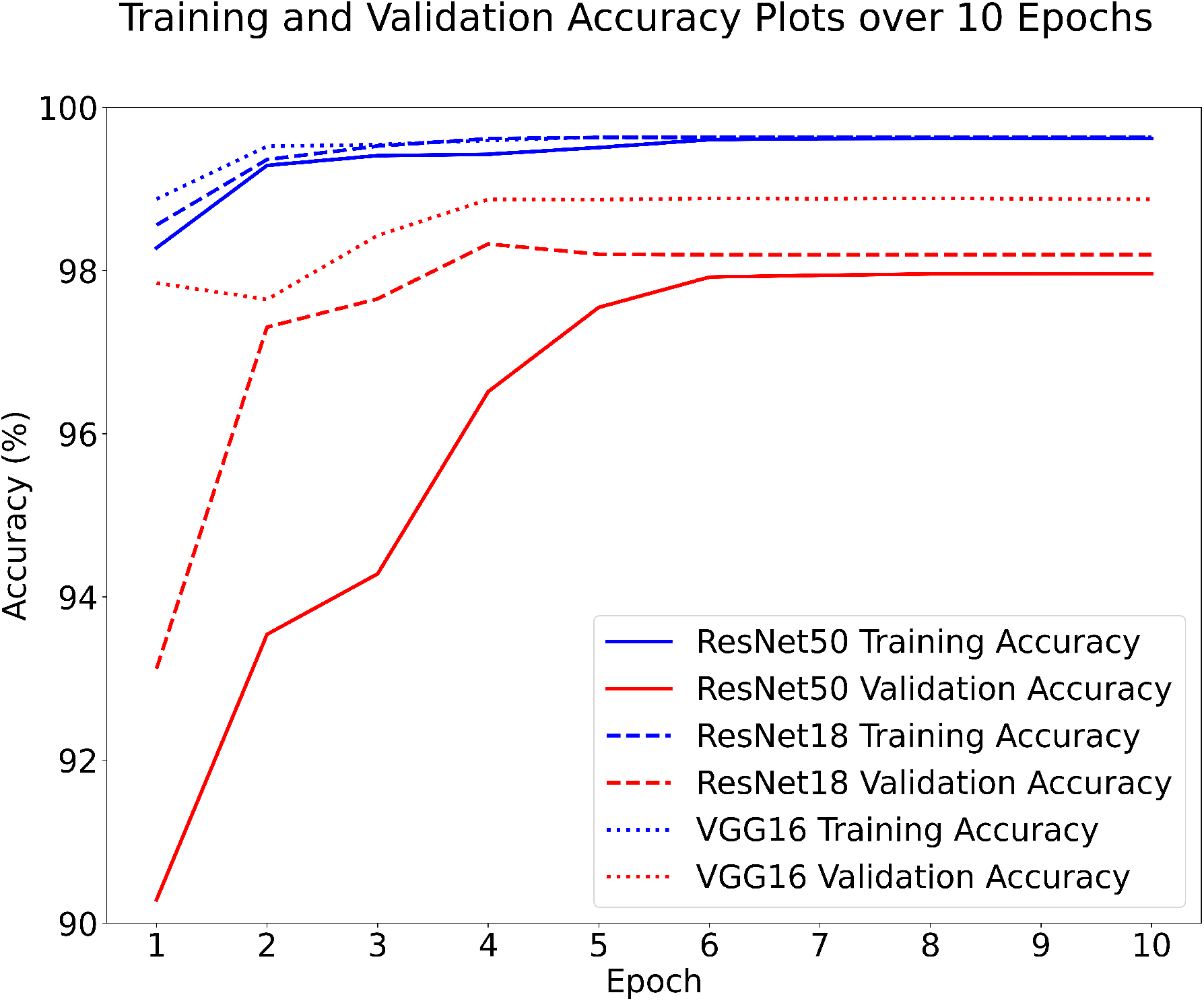
Training and validation accuracy each of the three networks, found at the end of each epoch. The small gap between the training and validation accuracies suggests that the models are not overfitting. Note the scale starts at 90%.

**Figure 5:**
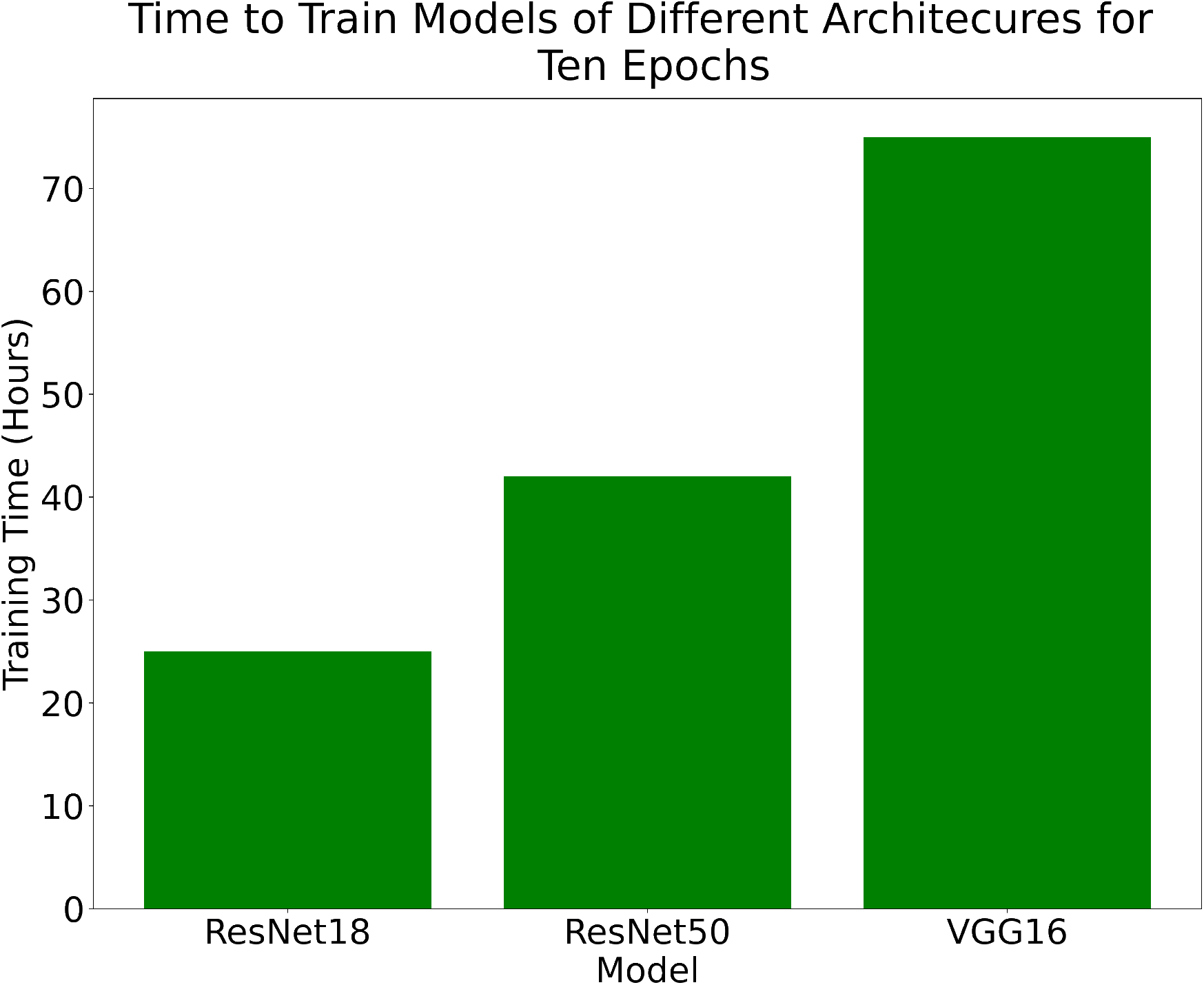
Time in hours to train the models for 10 epochs.The training and validation accuracy both level-off around epochs 5–6 which shows that the models are able to fit the data.

### 3.2 Test Set Accuracy

Figure 6 shows the accuracy of the three models. These results are in the high 90%’s across the train, validate and test sets which shows that the models have all learned the problem well. Table 2 shows the accuracy and balanced accuracy of each of the models on the test set. Figure 7 shows the confusion matrix for the ResNet18 model. The confusion matrix shows that the model performs very well on CT, MR and PET. Accuracy for X-rays can be improved by adding additional X-ray images across a larger spread of locations.

**Figure 6:**
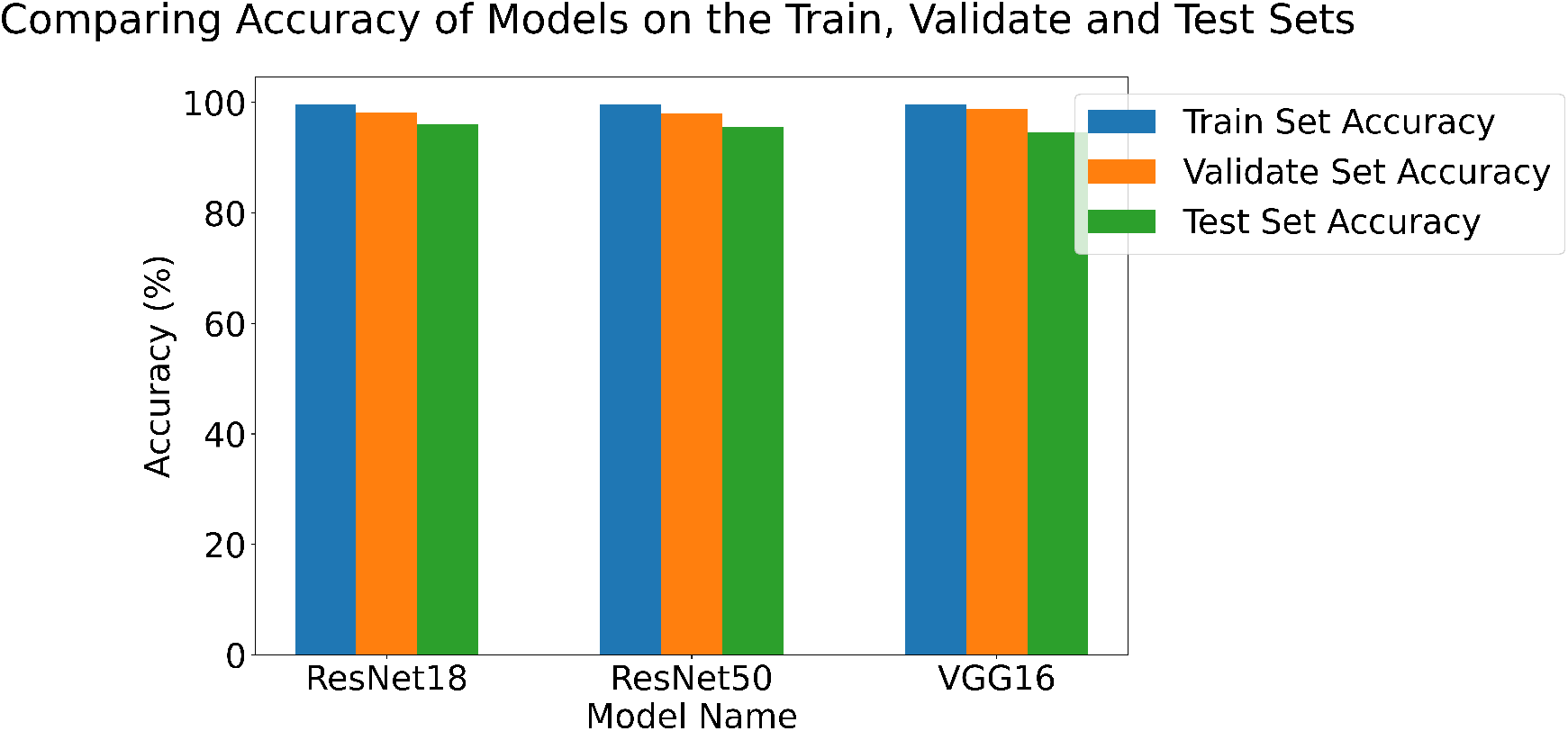
Accuracy of 3 models on the test set.

**Table 2:** Table containing the accuracy and balanced accuracy of various models on the test set. Each model was trained for 10 epochs.

| Model | Accuracy | Balanced Accuracy |
| --- | --- | --- |
| ResNet18 | 96.00% | 86.17% |
| ResNet50 | 95.60% | 85.65% |
| VGG16 | 94.58% | 81.08% |

**Figure 7:**
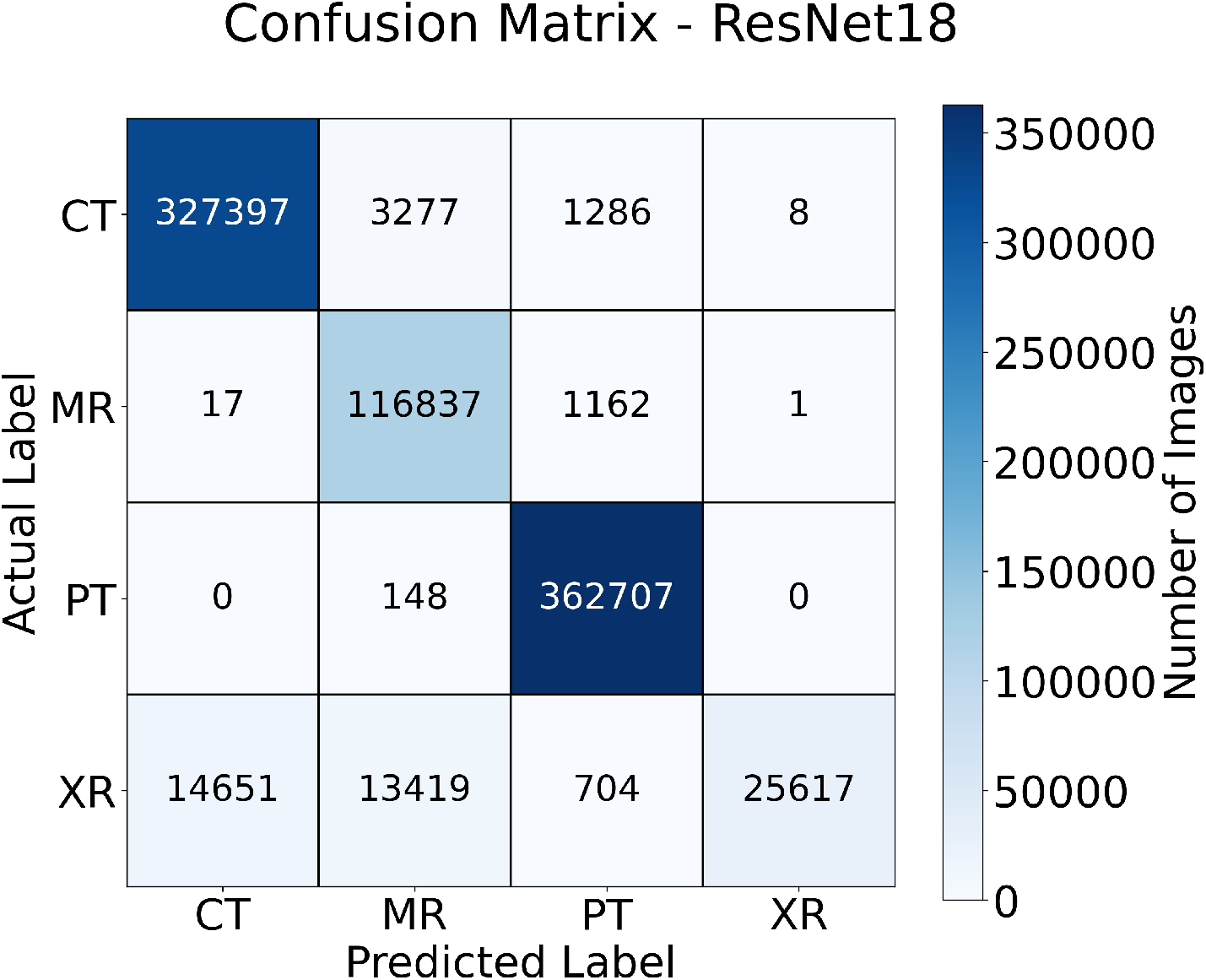
The confusion matrix for the ResNet18 on the test set. The model gains very high accuracy on the CT, MRI and PET. The ResNet18 results were chosen for this plot as this model achieved the highest accuracy and highest balanced accuracy.

### 3.3 Dataset Level Results

Table 3 shows the accuracy of the model on each dataset in the test set for the ResNet18 model, chosen because this model demonstrated superior classification performance over others tested in this study. It is interesting to note that in both tables the X-ray performance is in the 80-90% range for the Cancer Imaging Archive X-ray datasets, then drops for the MURA and Osteoarthritis Initiative datasets. This is likely because these datasets are bone X-rays, and most of the datasets only contain chest X-rays. Therefore, a better spread of X-ray datasets is needed for the performance of these models to be improved.

**Table 3:**
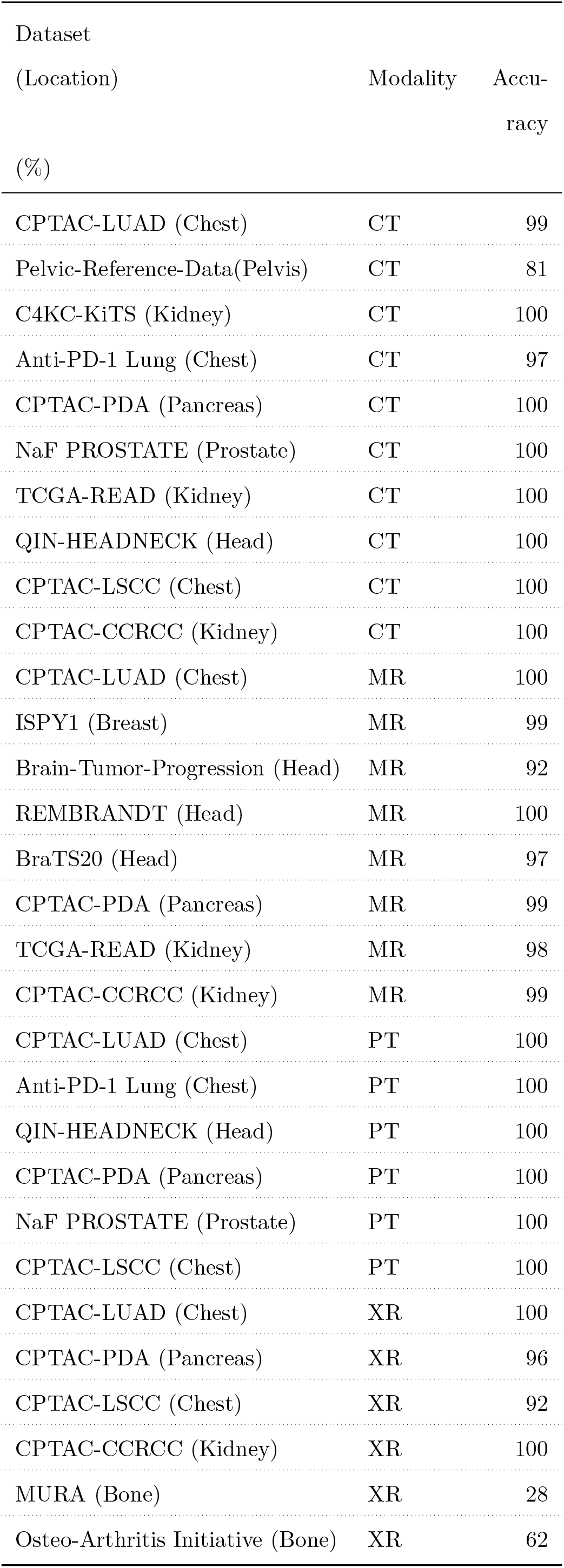
Table containing the accuracy of the ResNet18 model on every dataset in the test set. Some datasets appear more than once in this table because they contain multiple image modalities.

| Dataset<br>(Location) | Modality | Accuracy<br>(%) |
| --- | --- | --- |
| CPTAC-LUAD (Chest) | CT | 99 |
| Pelvic-Reference-Data(Pelvis) | CT | 81 |
| C4KC-KiTS (Kidney) | CT | 100 |
| Anti-PD-1 Lung (Chest) | CT | 97 |
| CPTAC-PDA (Pancreas) | CT | 100 |
| NaF PROSTATE (Prostate) | CT | 100 |
| TCGA-READ (Kidney) | CT | 100 |
| QIN-HEADNECK (Head) | CT | 100 |
| CPTAC-LSCC (Chest) | CT | 100 |
| CPTAC-CCRCC (Kidney) | CT | 100 |
| CPTAC-LUAD (Chest) | MR | 100 |
| ISPY1 (Breast) | MR | 99 |
| Brain-Tumor-Progression (Head) | MR | 92 |
| REMBRANDT (Head) | MR | 100 |
| BraTS20 (Head) | MR | 97 |
| CPTAC-PDA (Pancreas) | MR | 99 |
| TCGA-READ (Kidney) | MR | 98 |
| CPTAC-CCRCC (Kidney) | MR | 99 |
| CPTAC-LUAD (Chest) | PT | 100 |
| Anti-PD-1 Lung (Chest) | PT | 100 |
| QIN-HEADNECK (Head) | PT | 100 |
| CPTAC-PDA (Pancreas) | PT | 100 |
| NaF PROSTATE (Prostate) | PT | 100 |
| CPTAC-LSCC (Chest) | PT | 100 |
| CPTAC-LUAD (Chest) | XR | 100 |
| CPTAC-PDA (Pancreas) | XR | 96 |
| CPTAC-LSCC (Chest) | XR | 92 |
| CPTAC-CCRCC (Kidney) | XR | 100 |
| MURA (Bone) | XR | 28 |
| Osteo-Arthritis Initiative (Bone) | XR | 62 |

## 4 Conclusion

In this work, we proposed a hyper-scale classifier, capable of classifying diagnostic imaging data in the scale of millions of medical images, with significant classification accuracy. We used a dataset comprised of 4.5 million images to train a ResNet-50, ResNet-18, and VGG16 CNN. The trained classifiers were then tested for their classification accuracy on 4 modalities ((Computed Tomography (CT), Magnetic Resonance Imaging (MRI), Positron Emission Tomography (PET), and X-ray). The best performing model, among the ones tested demonstrated a classification accuracy of 96%. Our results show that hyper-scale classifiers are capable of accurately classifying volumes of image data encountered in real-word applications, such as those contained in image repositories or diagnostic imaging data collected by national healthcare institutions.

Future work on this topic will be to extend the scope of the hyper-scale modality classifier to work on 3D scan modalities, such as CT, MR, PET.

## Data Availability

The data used in this study comes from multiple sources, as indicated in Appendix Datasets. If not specified here then the data can be accessed with no special permission required by the data provider. Permission was required to access MURA, ChexPERT, MRNet, the NDA Osteo-Arthritis Initiative and BraTS20. Code created for this research is hosted on GitHub with an MIT Licence.

## A List of All Datasets Used

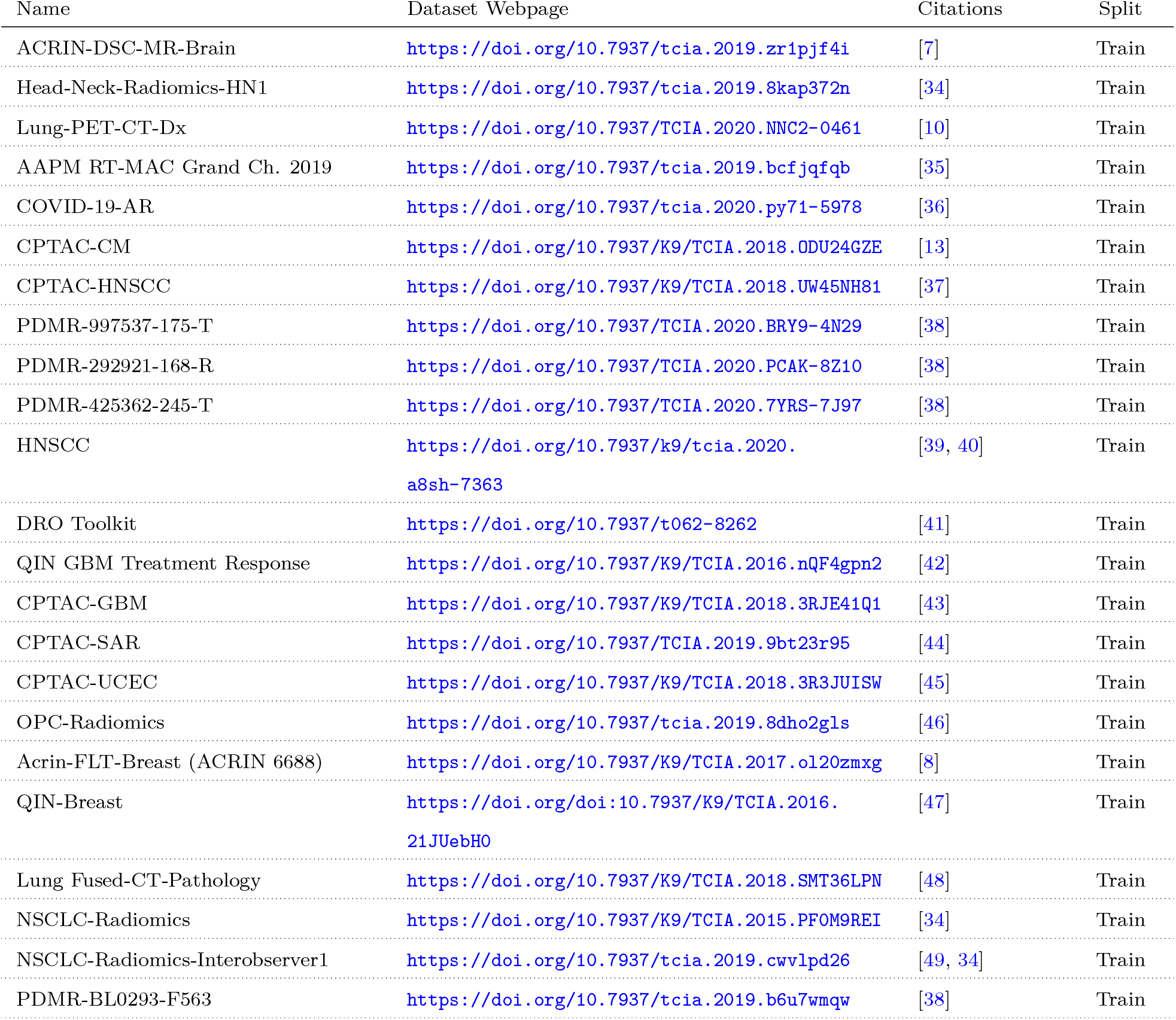
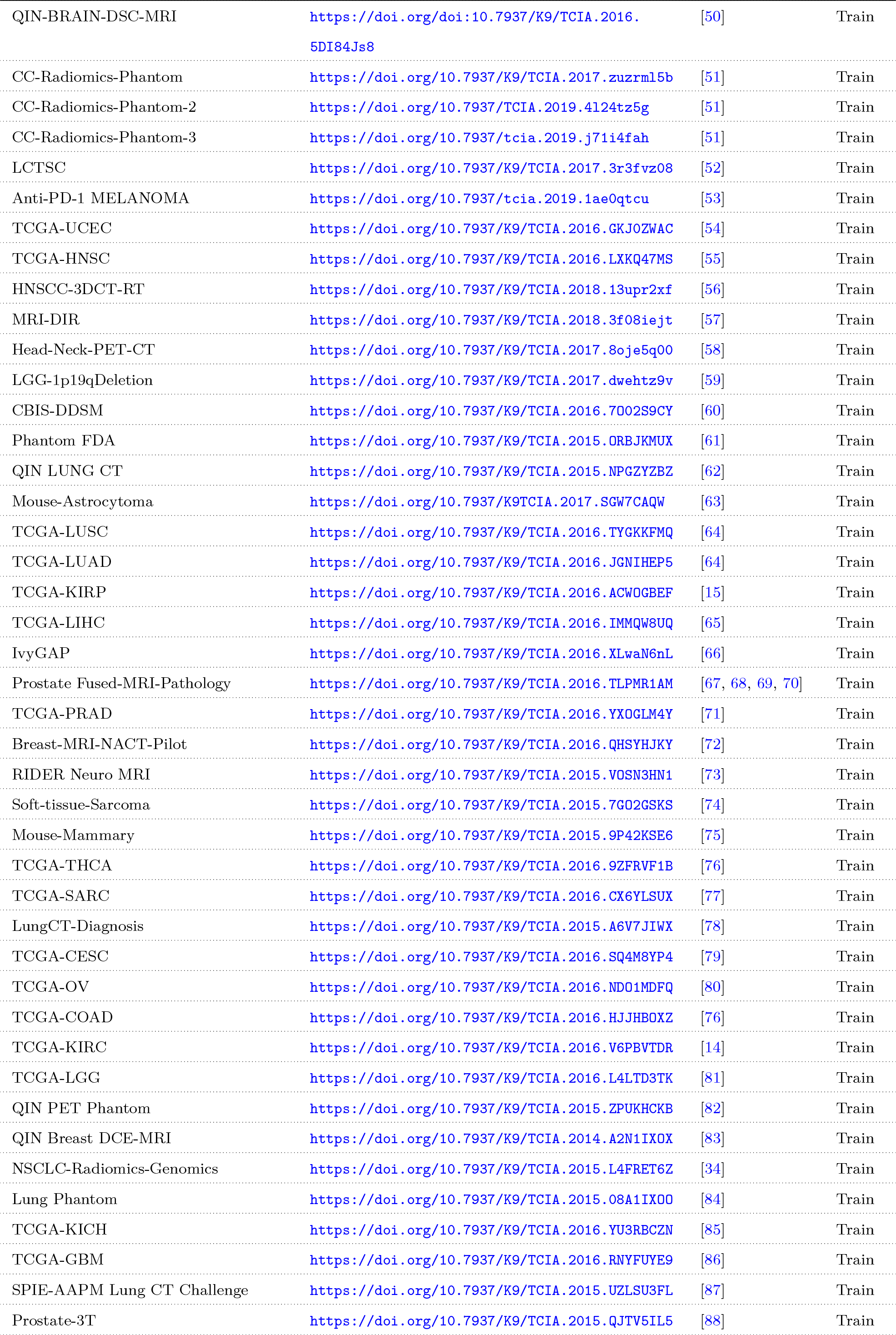
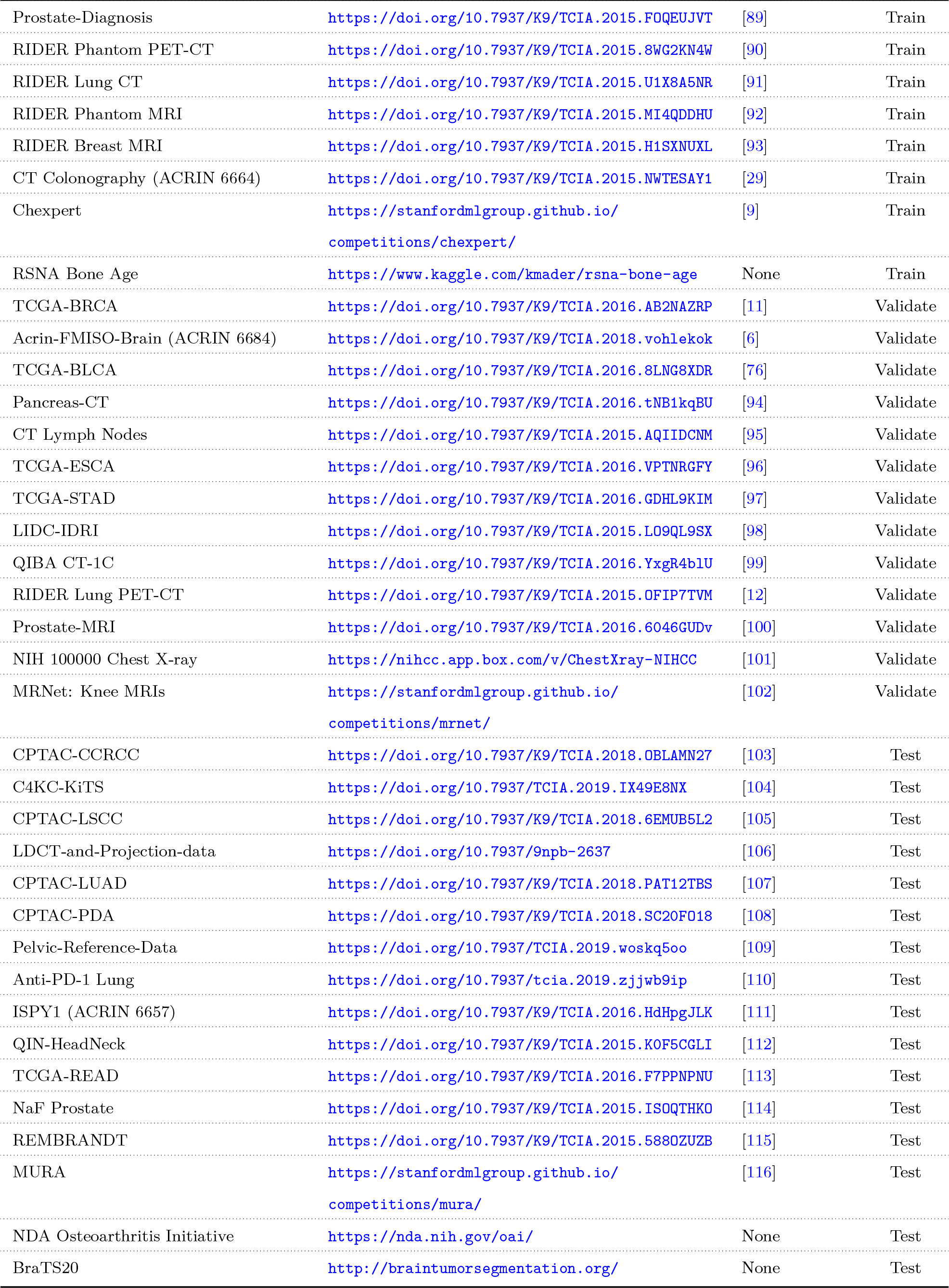

## Code, Data, & Materials Availability

The data used in this study comes from multiple sources, as indicated in Appendix A. If not specified here then the data can be accessed with no special permission required by the data provider. Permission was required to access MURA, ChexPERT, MRNet, the NDA Osteo-Arthritis Initiative and BraTS20.

Code created for this research is hosted on GitHub with an MIT Licence[33].

